# Loss of recognition of SARS-CoV-2 B.1.351 variant spike epitopes but overall preservation of T cell immunity

**DOI:** 10.1101/2021.06.03.21258307

**Authors:** Catherine Riou, Roanne Keeton, Thandeka Moyo-Gwete, Tandile Hermanus, Prudence Kgagudi, Richard Baguma, Houriiyah Tegally, Deelan Doolabh, Arash Iranzadeh, Lynn Tyers, Hygon Mutavhatsindi, Marius B. Tincho, Ntombi Benede, Gert Marais, Lionel R. Chinhoyi, Mathilda Mennen, Sango Skelem, Elsa du Bruyn, Cari Stek, SA-CIN, Tulio de Oliveira, Carolyn Williamson, Penny L. Moore, Robert J. Wilkinson, Ntobeko A. B. Ntusi, Wendy A. Burgers

**Affiliations:** Wellcome Centre for Infectious Diseases Research in Africa, University of Cape Town; Observatory 7925, South Africa; Institute of Infectious Disease and Molecular Medicine University of Cape Town; Observatory 7925, South Africa; Division of Medical Virology, Department of Pathology; University of Cape Town; Observatory 7925, South Africa; National Institute for Communicable Diseases of the National Health Laboratory Services; Johannesburg, South Africa; MRC Antibody Immunity Research Unit, School of Pathology, Faculty of Health Sciences, University of the Witwatersrand; Johannesburg, South Africa; KwaZulu-Natal Research Innovation and Sequencing Platform; Durban, South Africa; Department of Medicine, University of Cape Town and Groote Schuur Hospital; Observatory 7925, South Africa; Hatter Institute for Cardiovascular Research in Africa, Faculty of Health Sciences, University of Cape Town; Observatory 7925, South Africa; Department of Infectious Diseases, Imperial College London; W12 0NN, UK; The Francis Crick Institute; London, NW1 1AT, UK

## Abstract

SARS-CoV-2 variants have emerged that escape neutralization and potentially impact vaccine efficacy. T cell responses play a role in protection from reinfection and severe disease, but the potential for spike mutations to affect T cell immunity is poorly studied. We assessed both neutralizing antibody and T cell responses in 44 South African COVID-19 patients infected either with B.1.351, now dominant in South Africa, or infected prior to its emergence (‘first wave’), to provide an overall measure of immune evasion. We show for the first time that robust spike-specific CD4 and CD8 T cell responses were detectable in B.1.351-infected patients, similar to first wave patients. Using peptides spanning only the B.1.351 mutated regions, we identified CD4 T cell responses targeting the wild type peptides in 12/22 (54.5%) first wave patients, all of whom failed to recognize corresponding B.1.351-mutated peptides (p=0.0005). However, responses to the mutated regions formed only a small proportion (15.7%) of the overall CD4 response, and few patients (3/44) mounted CD8 responses that targeted the mutated regions. First wave patients showed a 12.7 fold reduction in plasma neutralization of B.1.351. This study shows that despite loss of recognition of immunodominant CD4 epitope(s), overall CD4 and CD8 T cell responses to B.1.351 are preserved. These observations may explain why, despite substantial loss of neutralizing antibody activity against B.1.351, several vaccines have retained the ability to protect against severe COVID-19 disease.

**One Sentence Summary:** T cell immunity to SARS-CoV-2 B.1.351 is preserved despite some loss of variant epitope recognition by CD4 T cells.

## INTRODUCTION

High levels of ongoing SARS-CoV-2 transmission have led to the emergence of new viral variants, which now dominate the pandemic. Variants of concern have been characterized as having increased transmissibility, potentially greater pathogenicity, and the ability to evade host immunity *(1)*. Four such variants of concern now circulate widely, namely B.1.1.7 in the US and Europe, B.1.351 in southern Africa, P.1 in Brazil and South America, and B.1.617 in India *(2–6)*. A primary concern is whether the immune response generated against ancestral SARS-CoV-2 strains, upon which all approved first generation vaccines are based, still confers protection against variants. The potential threat of reduced vaccine efficacy has prompted swift action from vaccine manufacturers, and an adapted mRNA vaccine based on B.1.351 has been developed and tested in clinical trials *(7)*.

SARS-CoV-2 variant B.1.351, which was first described in South Africa in October 2020*(5)*, is now responsible for >95% of infections in the country, and has spread across much of southern Africa *(6)*. It is the most concerning of the variants, tending to demonstrate the greatest reduction in neutralization sensitivity to COVID-19 convalescent and vaccinee plasma *(8–13)*, as well as reduced vaccine efficacy *(14–16)*. However, some vaccines have still demonstrated high efficacy against severe COVID-19 disease after B.1.351 infection *(16, 17)*, suggesting that T cell immunity plays an important role in immune protection, and may mitigate the effect of reduced neutralizing antibody activity.

To date, efforts to characterize immune evasion by SARS-CoV-2 variants have focused mainly on their ability to escape neutralization *(8–13)*. There is limited data addressing whether SARS-CoV-2 variants can evade T cell immunity *(18–22)* in natural infection or after vaccination. Furthermore, spike-specific T cell responses in COVID-19 patients infected with variant lineages have not been investigated. Here, we determined whether B.1.351 spike mutations affect the recognition of T cell epitopes in patients infected with the ancestral and B.1.351 SARS-CoV-2 lineages. We demonstrate for the first time that loss of CD4 T cell recognition does indeed occur in B.1.351-mutated spike regions, although the majority of the T cell response is maintained. Furthermore, B.1.351-infected patients mounted comparable spike responses as those infected with earlier strains. These results have important implications for reinfection and vaccine efficacy.

## RESULTS

### T cell responses in patients infected with ancestral strains or B.1.351

SARS-CoV-2 spike-specific neutralizing antibody and T cell responses were measured in hospitalized COVID-19 patients enrolled at Groote Schuur Hospital (Western Cape, South Africa) during the first wave of the COVID-19 pandemic (n = 22), prior to the emergence of the B.1.351 variant, and during the second wave of the pandemic (n = 22), after the B.1.351 variant became the dominant lineage (**Figure 1A**). During the first wave of COVID-19, 100% of sequenced virus corresponded to the ancestral SARS-CoV-2 lineages (Wuhan and D614G). Conversely, during the second wave in South Africa, the B.1.351 lineage accounted for >95% of reported SARS-CoV-2 infections at the time of sample collection (**Figure 1B)**. The B.1.351 variant is defined by nine amino acid changes in the spike protein, and all second wave participants that we sequenced (19/22) had confirmed infection with B.1.351, harboring 7 to 8 changes associated with the B.1.351 lineage *(5)* (**Figure S1**). Although SARS-CoV-2 viral sequences were not available for patients recruited in June to August 2020 during the first wave, we assumed that all participants were infected with the ancestral virus, since B.1.351 was first detected in October 2020 in the Western Cape.

**Fig. 1.**
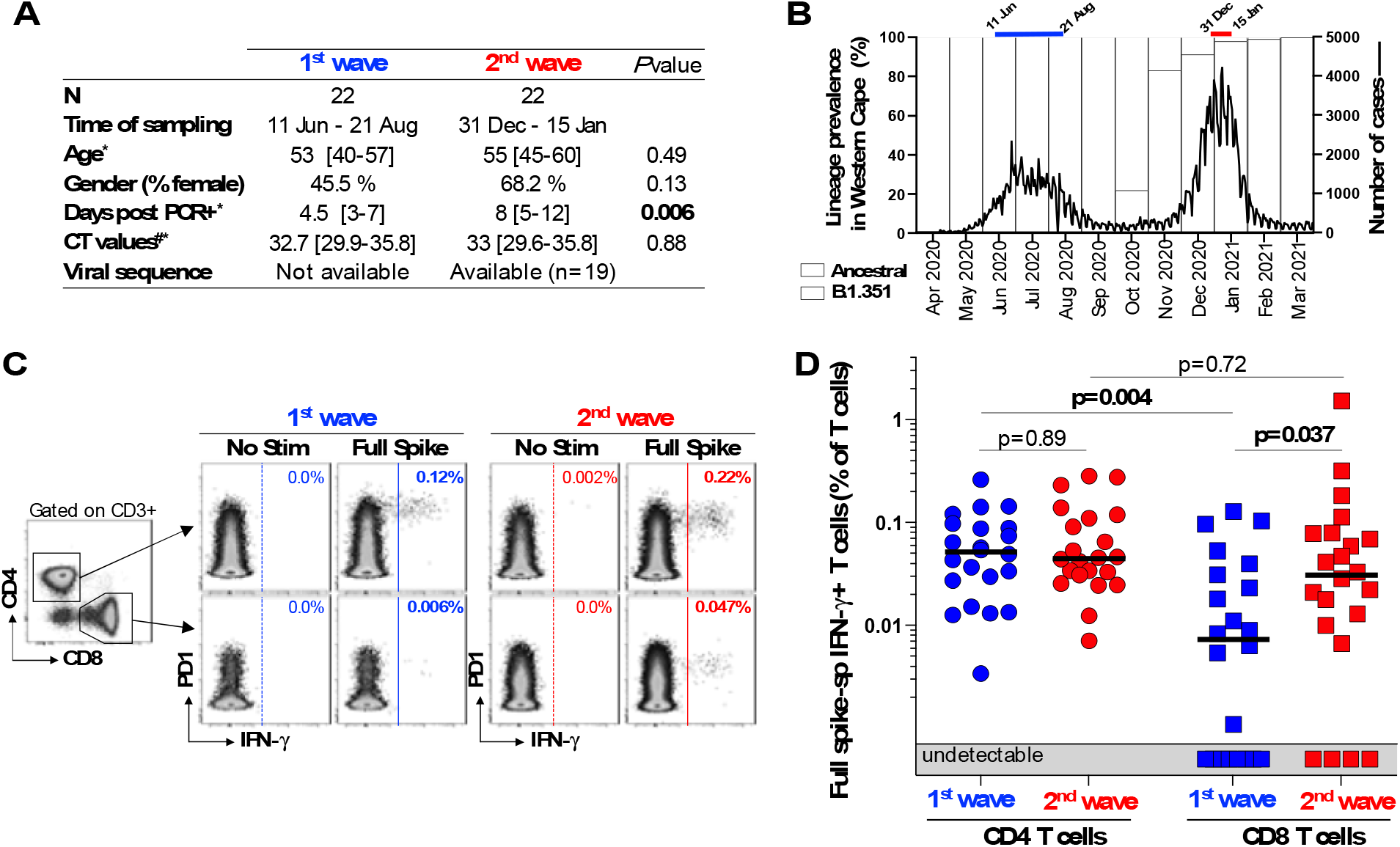
T cell recognition of SARS-CoV-2 spike in first and second wave COVID-19 patients. (**A**) Clinical characteristics of acute COVID-19 patients recruited during the first and second wave of the COVID-19 pandemic. ^*^: median and interquartile range. ^#^: SARS-CoV-2 polymerase chain reaction (PCR) was performed using the Allplex™ 2019-nCoV Assay (Seegene). The cycle threshold (CT) value for the N-gene is reported. (**B**) SARS-CoV-2 epidemiological dynamics in the Western Cape (South Africa). Prevalence of SARS-CoV-2 strains is reported on the left y-axis (based on the sequencing of 1178 samples). Ancestral strains are depicted in blue and B.1.351 strains (501Y.V2) in red. The number of COVID-19 cases is represented on the right y-axis. The bars on top of the graph indicate the periods when samples were collected in the first and second waves of the local epidemic. (**C**) Representative flow cytometry plots of IFN-γ production by CD4 (top panel) and CD8 (bottom panel) T cells in response to ancestral full spike peptide pool (Full Spike) in one first wave (blue) and one second wave (red) COVID-19 patient. Frequencies of IFN-γ-producing cells are indicated. (**D**) Summary graph of the frequency of SARS-CoV-2-specific CD4 or CD8 T cells in first wave (n = 22, blue) and second wave (n = 22, red) COVID-19 patients. Bars represent medians. Statistical analyses were performed using the Mann-Whitney test between first and second wave and the Wilcoxon test between CD4 and CD8 T cell responses.

First, we compared the magnitude of CD4 and CD8 T cell responses directed at the spike protein of SARS-CoV-2 in first and second wave patients. Using flow cytometry, we measured the production of IFN-γ in response to a peptide pool covering the full ancestral spike protein (‘Full spike’) (**Figure 1C**). All participants tested exhibited a CD4 response, with a comparable frequency of spike specific-CD4 T cells in first and second wave patients (0.051% and 0.045%, respectively, **Figure 1D**). As previously reported, the prevalence and magnitude of the SARS-CoV-2-specific CD8 T cell response was significantly lower compared to the CD4 response in the first wave *(23)*, with 63.6% (14/22) of first wave patients and 81.8% (18/22) of second wave patients exhibiting a detectable spike-specific CD8 T cell response. The median frequency of spike-specific CD8 T cells was higher in second wave patients compared to first wave patients (0.031 % and 0.007%, respectively, *P* = 0.037, **Figure 1D**). This may be explained by the fact that patients from the second wave were sampled at a later time post-PCR positivity compared to the patients recruited during the first wave (median: 8 days *vs*. 4.5 days, respectively, *P* = 0.006, **Figure 1A**). Overall, these data are in accordance with a recent report showing that T cell responses directed at the entire SARS-CoV-2 spike protein in convalescent COVID-19 donors were not substantially affected by mutations found in SARS-CoV-2 variants *(22)*.

### CD4 T cell targeting of variant spike epitopes

Since B.1.351-associated mutations occur only at a few residues of the spike protein, we then assessed the recognition of peptide pools selectively spanning the variable regions of spike, one composed of the ancestral version of the peptides (‘WT pool’) and the other composed of B.1.351-mutated peptides (‘B.1.351 pool’) (**Table S1**). In patients recruited during the first wave, IFN-γ CD4 T cell responses to the WT pool were detectable in 54.5% (12/22) patients (**Figure 2A-B**). In those who mounted responses, the magnitude of the WT pool response was ∼ 6.4-fold lower than full spike responses (median: 0.0075% vs 0.048%, respectively, *P* < 0.0001). In the 12 participants responding to the WT pool, the overall median relative contribution of WT epitopes located at spike mutation sites to the total spike-specific CD4 T cell response was 15.7%, ranging from 5.7% to 24%. These results suggest that the majority of SARS-CoV-2 spike specific-CD4 T cell responses are directed against conserved epitopes between the ancestral and the B.1.351 lineage. When we tested the corresponding B.1.351 pool, all 12 of the first wave WT pool responders failed to cross-react with the mutated peptides from B.1.351 (**Figure 2B**, left panel). These results show that B.1.351 spike mutated epitopes were no longer recognized by CD4 T cells targeting the WT epitopes, demonstrating that this loss of recognition by CD4 T cells is likely mediated by variant mutations. This is broadly consistent with recent data from mRNA vaccinees, where full spike pools containing B.1.351 mutated peptides detected T cell responses that were diminished by 30% compared to ancestral full spike, revealing that the mutated sequences mediate differential recognition but make up a minor contribution to the overall SARS-CoV-2 spike specific-T cell response *(20)*.

**Fig. 2.**
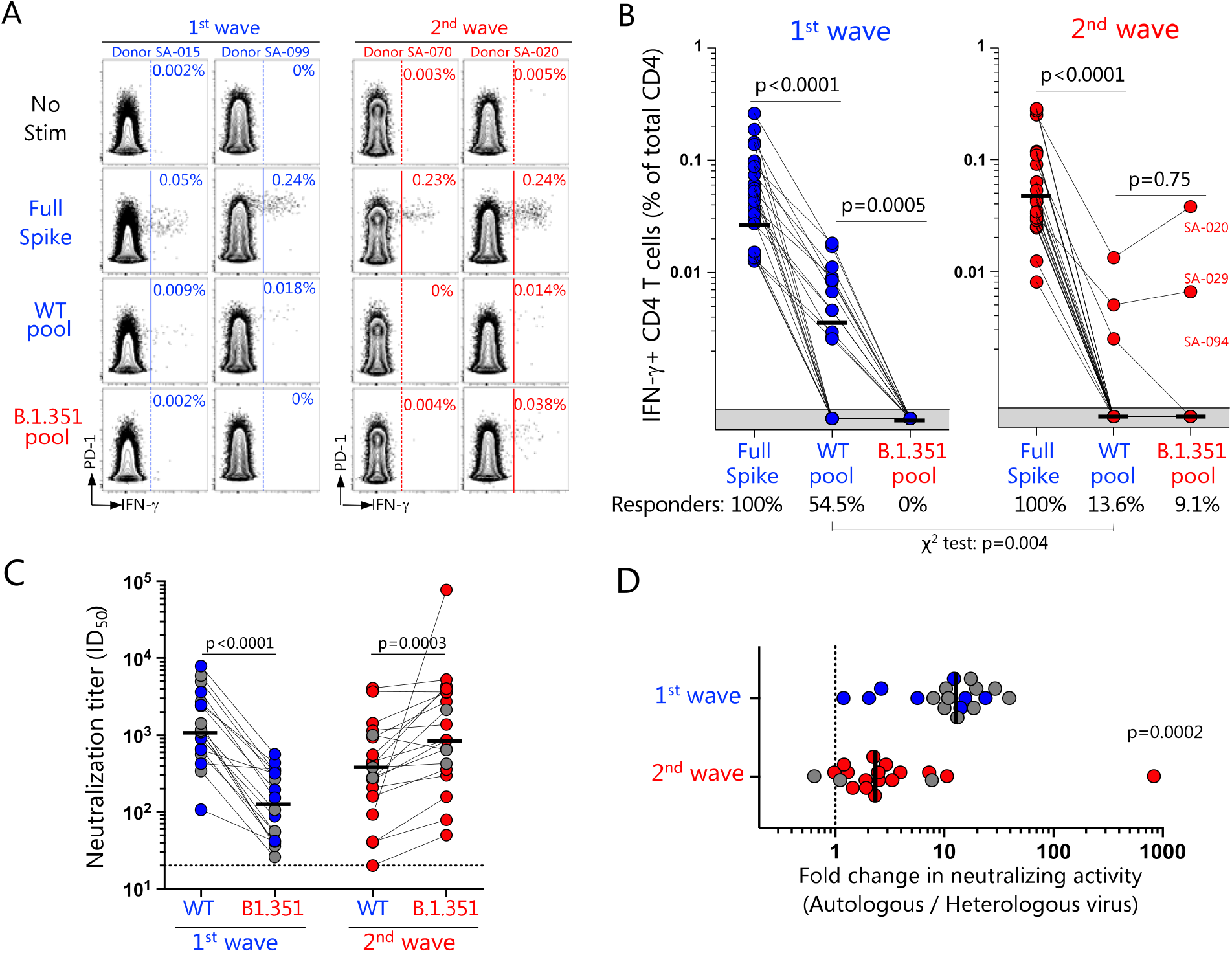
Loss of recognition of SARS-CoV-2 B.1.351 variant epitopes and neutralizing antibody responses. (**A**) Representative flow cytometry plots of IFN-γ production by CD4 T cells in response to ancestral full spike peptide pool (Full spike), and smaller pools selectively spanning the mutated regions of ancestral spike (WT pool) or B.1.351 spike (B.1.351 pool) in two first wave (blue) and two second wave (red) COVID-19 patients. Frequencies (%) of IFN-γ positive cells are indicated on the plots. (**B**) Summary graphs of the frequency of IFN-γ-producing SARS-CoV-2-specific CD4 T cells in first wave (n = 22, left) and second wave (n = 22, right) COVID-19 patients. The proportion of patients exhibiting a detectable response to the different peptide pools (i.e., responders) is indicated at the bottom of each graph. (**C**) Plasma samples from COVID-19 patients recruited during the first wave (n = 18) or the second wave (n = 19) were tested for their neutralization cross-reactivity against the ancestral or B1.351 pseudoviruses. The threshold of detection for the neutralization assay was a 50% inhibitory dilution (ID_50_) of 20. Gray dots indicate patients who displayed a detectable CD4 T cell response to the WT pool, selectively covering the variable regions of spike, and lost recognition to the B.1.351 pool. Neutralization data on the second wave cohort are from *(25)*. (**D**) Fold-change in neutralization titers is shown for the data in **c**. Bars represent medians. Statistical analyses were performed using the Wilcoxon test and the chi-squared test.

We next measured peptide responses in patients infected with the B.1.351 lineage. The B.1.351 pool was not readily recognized by patients infected with the homologous variant (2/22; 9.1%) (**Figure 2B**, right panel). A single donor had a detectable response to the WT but not the B.1.351 pool. These data suggest that mutations in B.1.351 spike epitopes abrogate epitope immunogenicity by altering their processing and/or presentation, consistent with the loss of recognition of B.1.351 mutated peptides by T cells in first wave patients. The few responders may represent individuals with uncommon HLA alleles able to present peptides within the mutated pool. Further analysis will be necessary to define the specific epitope(s) and presenting HLA alleles accounting for these CD4 responses.

In order to obtain an overall measure of immune escape in our participants, we measured their neutralizing antibody responses to the ancestral and B.1.351 spike protein, using a pseudotyped lentivirus neutralization assay (**Figure 2C-D**). As we showed previously *(11)* in patients infected with the ancestral strains (first wave), a considerable loss of neutralization activity was observed against B.1.351 (median fold change: 12.7, IQR: 7.3-18.8). In contrast, patients infected with B.1.351 (second wave) retained a substantial capacity to neutralize the ancestral virus, as shown by a moderate reduction in neutralizing activity against the ancestral strain (median: 2.3, IQR: 1.3-3.9). Of note, in the six first wave patients where loss of cross-neutralization was profound (titer <100), it is reassuring that the T cell response was relatively intact, thus providing some cross-protection. We found no association between the frequency of SARS-CoV-2 spike-specific CD4 T cell responses and neutralizing activity (data not shown), consistent with an earlier study *(24)*.

### CD8 T cell targeting of variant spike epitopes

Finally, we also defined the recognition of WT and B.1.351 peptide pools by CD8 T cells in both patient groups (**Figure 3**). Regardless of the infecting SARS-CoV-2 lineage, peptides covering the spike mutation sites were rarely recognized by CD8 T cells, with only 3/44 (6.8%) of patients exhibiting a CD8 response, one in the first wave cohort and two in the second wave cohort. Thus, in contrast to CD4 T cells, the regions in which B.1.351 mutations occur are not commonly targeted by CD8 T cells. Moreover, in the three patients with a CD8 response, the frequency of IFN-g producing CD8 T cells was comparable between the WT and the B.1.351 pool stimulation, indicating that mutations did not affect epitope recognition (**Figure 3B**). Overall, these data indicate that B.1.351 mutations do not affect CD8 T cell responses in these experiments.

**Fig. 3.**
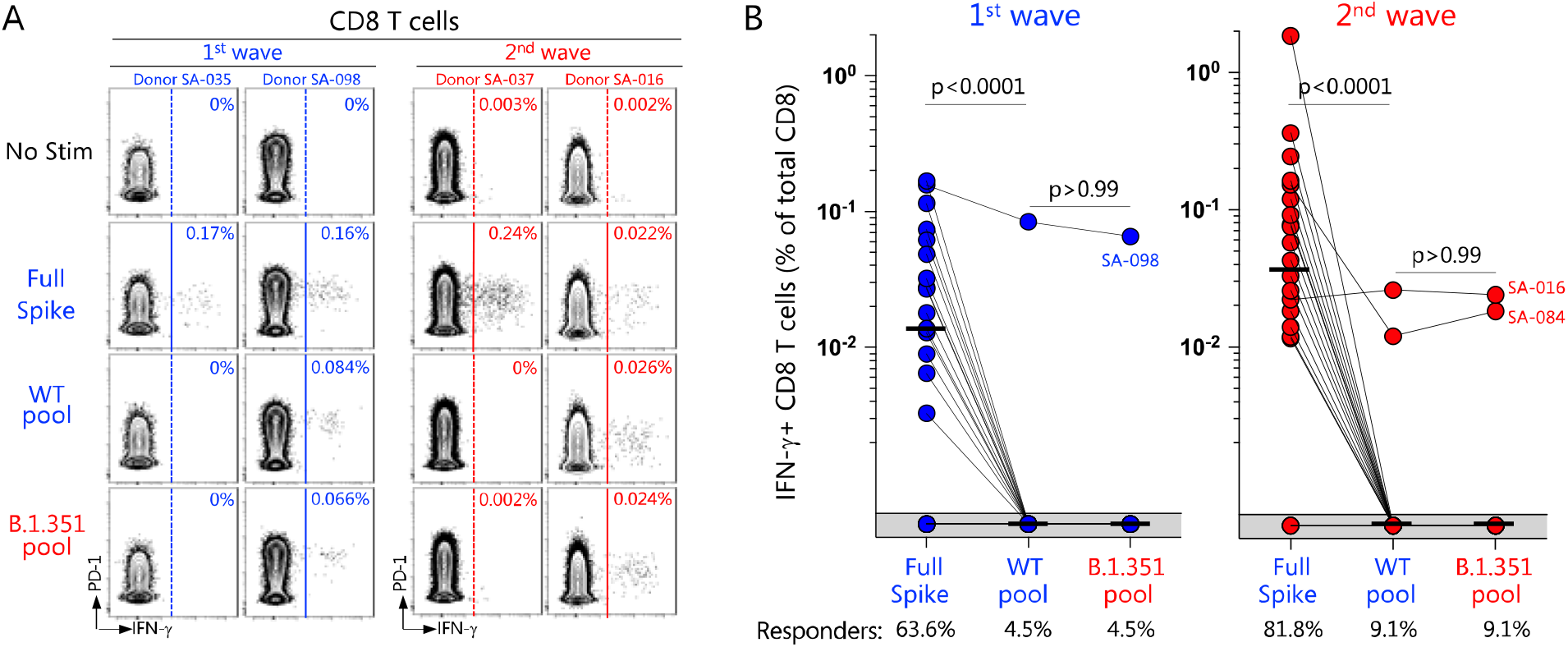
Infrequent recognition of SARS-CoV-2 ancestral or B.1.351 variant spike epitopes by CD8 T cells. (**A**) Representative flow cytometry plots of IFN-γ production by CD8 T cells in response to ancestral full spike peptide pool (Full Spike), and pools covering the mutated regions of ancestral spike (WT pool) or B.1.351 spike (B.1.351 pool) in two first wave (blue) and two second wave (red) COVID-19 patients. Frequencies (%) of IFN-γ positive cells are indicated on the plots. (**B**) Summary graphs of the frequency of IFN-γ-producing SARS-CoV-2-specific CD8 T cells in first wave (n = 22, left) and second wave (n = 22, right) COVID-19 patients. The proportion of patients exhibiting a detectable response to the different peptide pools (i.e. responders) is indicated at the bottom of each graph. Bars represent medians. Statistical analyses were performed using the Wilcoxon test.

## DISCUSSION

In this study, we demonstrate that infection with the B.1.351 variant results in robust T cell responses, comparable to responses elicited to ancestral strains. This work extends our recent findings on neutralizing antibody responses elicited by B.1.351 *(25)*. We also demonstrate for the first time that the recognition of epitopes by CD4 T cells targeting variable spike regions was affected by B.1.351 spike mutations in patients infected with ancestral lineages. However, it is reassuring that the loss of recognition of B.1.351 mutated spike epitopes has a minor impact on the overall CD4 Th1 cell response. Moreover, the CD8 T cell response to spike was unaffected.

In contrast to neutralizing antibody epitopes, T cell epitopes are located along the full length of the spike protein and T cell responses are broadly targeted in natural infection *(22, 26– 29)*. Thus, it is unsurprising that the B.1.351 variant retains the ability to generate strong T cell immune responses, as B.1.351 spike mutations are limited to a few residues. The immunodominance of ancestral epitope(s) that appear to be all but lost in B.1.351 infection (52% vs 8% responders) is consistent with these mutated epitopes no longer being processed or presented in B.1.351 infection, thus not eliciting a response. Altered epitope hierarchies and the presentation of subdominant epitopes *(30)* may further explain the strong B.1.351 full spike responses. Although it remains to be determined whether vaccination generates broad spike responses similar to natural infection, it appears unlikely that vaccine-induced T cell immunity will fail to recognize SARS-CoV-2 variants.

Viral evasion of cytotoxic T lymphocyte or T helper recognition may result in delayed clearance of infected cells, or inadequate help provided to B cells, influencing the antibody response. Viral escape from specific SARS-CoV-2 CD8 epitopes has recently been described, in spike, nucleocapsid and ORF3a proteins *(18, 19, 21)*. Whilst CD8 T cells can exert selective pressure on viruses resulting in mutational escape, CD4 T helper responses act indirectly, and it is not clear how they could drive viral evolution. Mutations occurring in response to immune pressure from neutralizing antibodies, or associated with increased viral infectivity or greater protein stability *(21)* could coincide with CD4 epitopes, and thus represent ‘collateral damage’ for the CD4 response.

Further work in a greater number of participants may identify additional responders to epitopes in the mutated regions of spike, given polymorphism in HLA genes. It remains to be determined which specific epitopes within spike variable regions are immunogenic. A number of shared mutations are found in several variants of concern or interest, having arisen through convergent evolution *(31)*. Our findings would have generalizability beyond the B.1.351 variant if loss of recognition occurred in mutated regions that occurred in multiple variants. Examining responses to B.1.351 in the context of full mutated spike *(20, 22)* would corroborate our findings regarding the degree to which the overall spike T cell response is affected by mutations.

In conclusion, these results advance our understanding of cross-reactive T cell immunity in the context of viral variability, and highlight the importance of monitoring both antibody and T cell responses to emerging SARS-CoV-2 variants. We demonstrate a limited effect of viral mutations on T cell immunity which may explain why, despite substantial loss of neutralizing antibody activity against B.1.351, some vaccines have retained the ability to protect against severe COVID-19 disease. Whilst second generation vaccines based on SARS-CoV-2 variants are desirable, they may not be needed to generate improved T cell responses, and therefore the rollout of present vaccines must continue apace.

## MATERIALS AND METHODS

### Study Design

Hospitalized patients with PCR-confirmed acute COVID-19 were enrolled at Groote Schuur Hospital (Cape Town, Western Cape, South Africa) between June 11^th^ and August 21^st^, 2020 (first wave, n = 22) and between December 31^st^, 2020 and January 15^th^, 2021 (second wave, n = 22). The clinical characteristics of participants are summarized in **Figure 1A**. Blood samples were obtained at a median of 4.5 days [interquartile range (IQR): 3-7] after a positive PCR test for SARS-CoV-2 for patients from the first wave, and 8 days [IQR: 4-16] for second wave patients. Spike viral sequences were available for 19 of the 22 participants recruited during the second wave. Details on the spike sequence for each of these participants are presented in **Figure S1**. T cell immune responses were assessed by stimulating PBMC with peptide pools spanning full-length spike or smaller pools covering the regions mutated in B.1.351, followed by intracellular cytokine staining and flow cytometry (**Figure S2**). The study was approved by the University of Cape Town Human Research Ethics Committee (HREC: 207/2020 and R021/2020) and electronic or written informed consent was obtained from all participants.

### SARS-CoV-2 spike whole genome sequencing

Whole genome sequencing of SARS-CoV-2 was performed using nasopharyngeal swabs obtained from 19 of the hospitalized patients recruited during the second COVID-19 wave. Sequencing was performed as previously published *(25)*. Briefly, cDNA was synthesized from RNA extracted from the nasopharyngeal swabs using the Superscript IV First Strand synthesis system (Life Technologies, Carlsbad, CA) and random hexamer primers. Whole genome amplification was then performed by multiplex PCR using the ARTIC V3 protocol (https://www.protocols.io/view/ncov-2019-sequencing-protocol-v3-locost-bh42j8ye). PCR products were purified with AMPure XP magnetic beads (Beckman Coulter, CA) and quantified using the Qubit dsDNA High Sensitivity assay on the Qubit 3.0 instrument (Life Technologies Carlsbad, CA). The Illumina® DNA Prep kit was used to prepare indexed paired end libraries of genomic DNA. Sequencing libraries were normalized to 4 nM, pooled, and denatured with 0.2 N sodium hydroxide. Libraries were sequenced on the Illumina MiSeq instrument (Illumina, San Diego, CA, USA). The quality control checks on raw sequence data and the genome assembly were performed using Genome Detective 1.132 (https://www.genomedetective.com) and the Coronavirus Typing Tool *(32)*. The initial assembly obtained from Genome Detective was polished by aligning mapped reads to the references and filtering out low-quality mutations using bcftools 1.7-2 mpileup method. Mutations were confirmed visually with bam files using Geneious software (Biomatters Ltd, New Zealand). Phylogenetic clade classification of the genomes in this study consisted of analyzing them against a global reference dataset using a custom pipeline based on a local version of NextStrain (https://github.com/nextstrain/ncov) *(33)*. The workflow performs alignment of genomes, phylogenetic tree inference, tree dating and ancestral state construction and annotation. The phylogenetic trees were visualized using ggplot and ggtree *(34)*.

### Ancestral (wild type) and B.1.351 variant SARS-CoV-2 spike peptides

To assess the overall response to the full length SARS-CoV-2 spike protein, we combined two commercially available peptide pools (PepTivator®, Miltenyi Biotech, Bergisch Gladbach, Germany) including: i) a pool of peptides (15-mer sequences with 11 amino acids (aa) overlap) covering the ancestral N-terminal S1 domain of SARS-CoV-2 (GenBank MN908947.3, Protein QHD43416.1) from aa 1 to 692 and ii) a pool of peptides (15-mer sequences with 11 aa overlap) covering the immunodominant sequence domains of the ancestral C-terminal S2 domain of SARS-CoV-2 (GenBank MN908947.3, Protein QHD43416.1) including the sequence domains aa 683-707, aa 741-770, aa 785-802, and aa 885-1273. These pools were resuspended in distilled water at a concentration of 50 µg/mL. Individual peptides (15-mer sequences with 10 aa overlap) spanning ancestral or B.1.351 spike mutation sites (L18F, D80A, D215G, del 242-244, R246I, K417N, E484K, N501Y and A701V) were synthesized (GenScript Biotech, Piscataway, NJ, USA) and individually resuspended in dimethyl sulfoxide (DMSO; Sigma-Aldrich, St. Louis, Missouri, United States) at a concentration of 20 µg/mL. The peptide sequences are provided in **Table S1**, which also indicates where their recognition has been previously described *(22, 27, 29)*. Ancestral or B.1.351 pools (16 peptides) selectively spanning the mutated regions were created by pooling aliquots of these individual peptides at a final concentration of 160 µg/mL.

### Isolation of peripheral blood mononuclear cells (PBMC)

Blood was collected in heparin tubes and processed within 3 hours of collection. Peripheral blood mononuclear cells (PBMC) were isolated by density gradient sedimentation using Ficoll-Paque (Amersham Biosciences, Little Chalfont, UK) as per the manufacturer’s instructions and cryopreserved in freezing media consisting of heat-inactivated fetal bovine serum (FBS, Thermo Fisher Scientific, Oslo, Norway) containing 10% DMSO and stored in liquid nitrogen until use.

### Cell stimulation and flow cytometry staining

Cryopreserved PBMC were thawed, washed and rested in RPMI 1640 containing 10% heat-inactivated FCS for 4 hours prior to stimulation. PBMC were seeded in a 96-well V-bottom plate at ∼2 × 10^6^ PBMC per well and stimulated with SARS-CoV-2 spike peptide pools: full spike pool (Miltenyi), and ancestral and B.1.351 pools selectively spanning the mutated regions (4 µg/mL). All stimulations were performed in the presence of Brefeldin A (10 µg/mL, Sigma-Aldrich, St Louis, MO, USA) and co-stimulatory antibodies against CD28 (clone 28.2) and CD49d (clone L25) (1 µg/mL each; BD Biosciences, San Jose, CA, USA). As a negative control, PBMC were incubated with co-stimulatory antibodies, Brefeldin A and an equimolar amount of DMSO.

After 16 hours of stimulation, cells were washed, stained with LIVE/DEAD™ Fixable Near-IR Stain (Invitrogen, Carlsbad, CA, USA) and subsequently surface stained with the following antibodies: CD4 BV785 (OKT4, Biolegend, San Diego, CA, USA), CD8 BV510 (RPA-8, Biolegend), PD-1 PE (J105, eBioscience, San Diego, CA, USA). Cells were then fixed and permeabilized using a Transcription Factor Fixation buffer (eBioscience) and stained with CD3 BV650 (OKT3), IFN-g BV711 (4S.B3) from Biolegend. Finally, cells were washed and fixed in 1% formaldehyde in PBS. Samples were acquired on a BD LSR-II flow cytometer and analyzed using FlowJo (v9.9.6, FlowJo LLC, Ashland, OR, USA). A cytokine response was defined as positive when the frequency of cytokine produced in stimulated wells was at least twice the background of unstimulated cells. For the data presented, the background has been subtracted.

### SARS-CoV-2 pseudovirus based neutralization assay

SARS-CoV-2 pseudotyped lentiviruses were prepared by co-transfecting the HEK 293T cell line with the SARS-CoV-2 614G spike (D614G) or SARS-CoV-2 501Y.V2 spike (L18F, D80A, D215G, K417N, E484K, N501Y, A701V, 242-244 del) plasmids with a firefly luciferase encoding lentivirus backbone plasmid. The parental plasmids were kindly provided by Drs Elise Landais and Devin Sok (IAVI). For the neutralization assays, heat-inactivated plasma samples were incubated with the SARS-CoV-2 pseudotyped virus for 1 hour at 37°C, 5% CO_2_. Subsequently, 1×10^4^ HEK293T cells engineered to over-express ACE-2, kindly provided by Dr Michael Farzan (Scripps Research Institute), were added and the incubated at 37°C, 5% CO_2_ for 72 hours, upon which the luminescence of the luciferase gene was measured. CB6 and CA1monoclonal antibodies were used as controls.

### Statistical analyses

Analyses were performed in Prism (v9; GraphPad Software Inc, San Diego, CA, USA). Non-parametric tests were used for all comparisons. The Mann-Whitney and Wilcoxon tests were used for unmatched and paired samples, respectively. *P* values less than 0.05 were considered to indicate statistical significance.

## Data Availability

The data that support the findings of this study are available from the corresponding authors, WAB and CR, upon reasonable request.

## Supplementary Materials

### Supplementary figures

Fig. S1. Genomic sequencing confirmation of SARS-CoV-2 B.1.351 infection of COVID-19 second wave patients.

Fig. S2. Graphical representation of study approach.

### Supplementary tables

Table S1. Peptides included in the ancestral and B.1.351 peptide pools.

## Acknowledgments

The authors thank the study participants and their families, and the clinical staff and personnel at Groote Schuur Hospital in Cape Town for their support and dedication. We thank the informal 501Y.V2 consortium of South African scientists, chaired by Drs Willem Hanekom and Tulio de Oliveira, for suggestions and discussion of data.

## Funding

South African Medical Research Council

Wellcome Centre for Infectious Diseases Research in Africa (CIDRI-Africa), supported by the Wellcome Trust 203135/Z/16/Z and 222754

EDCTP2 programme of the European Union (EU)’s Horizon 2020 programme grants

TMA2017SF-1951-TB-SPEC and TMA2016SF-1535-CaTCH-22 (CR, WAB).

National Institutes of Health grant R21AI148027 (CR)

South African Research Chairs Initiative of the Department of Science and Innovation and the National Research Foundation grant 9834 (PLM)

National Research Foundation Postdoctoral Fellowship 129614 (HM)

Francis Crick Institute which receives funding from Wellcome FC0010218, UKRI FC0010218 and CRUK FC0010218 (RJW)

Rosetrees Trust grant M926 (CR, RJW)

For the purposes of open access the authors have applied a CC BY public copyright license to any author-accepted version arising from this submission.

## Author contributions

Conceptualization: CR, WAB, RJW, PLM, NABN

Investigation: CR, RK, RB, HM, MBT, NB, TH, TM-G, PK, HT, DD, LT, AI, GM, MM, LRC, SS, EdB, CS

Funding acquisition: WAB, CR, RJW, NABN

Supervision: CR, WAB, PLM, CW, TdO, RJW, NABN

Writing – original draft: CR,WAB

Writing – review & editing: All authors

## Competing interests

Authors declare that they have no competing interests.

## Data and materials availability

All data are available in the main text or the supplementary materials.

## Notes

### Competing Interest Statement

The authors have declared no competing interest.

### Funding Statement

We acknowledge funding from the South African Medical Research Council and the Wellcome Centre for Infectious Diseases Research in Africa (CIDRI-Africa) which is supported by core funding from the Wellcome Trust [203135/Z/16/Z and 222754]. CR and WAB are supported by the EDCTP2 programme of the European Union (EU) Horizon 2020 programme (TMA2017SF-1951-TB-SPEC to CR and TMA2016SF-1535-CaTCH-22 to WAB). CR is further supported by the National Institutes of Health (NIH) (R21AI148027). PLM is supported by the South African Research Chairs Initiative of the Department of Science and Innovation and the National Research Foundation (Grant No 9834). HM is supported by a National Research Foundation Postdoctoral Fellowship (Grant No 129614). RJW is supported by Francis Crick Institute which receives funding from Wellcome (FC0010218), UKRI (FC0010218) and CRUK (FC0010218). CR and RJW also receive support from Rosetrees Trust (M926). The authors or their institutions did not at any time receive payment or services from any other third party for any aspect of the submitted work.

### Author Declarations

The study was approved by the University of Cape Town Human Research Ethics Committee (HREC: 207/2020 and R021/2020) and electronic or written informed consent was obtained from all participants.

