## Supplementary materials for "Loss of recognition of SARS-CoV-2 B.1.351 variant spike epitopes but overall preservation of T cell immunity"

for

[†]South African Cellular Immunology Network (SA-CIN) collaborators listed at the end of this document

**This PDF file contains:**

Fig. S1
Fig. S2
Table S1
SA-CIN (South African Cellular Immunology Network) collaborator list

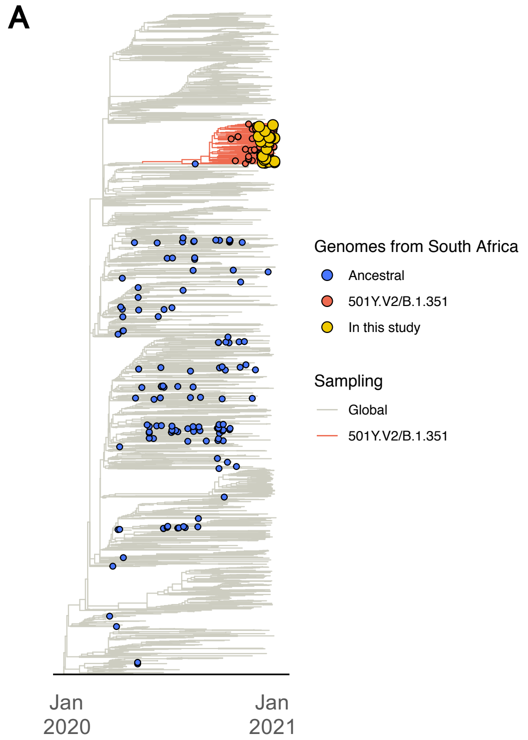


B

| | Position & amino acid | | | | | | | | |
|---|---|---|---|---|---|---|---|---|---|
| | 18 | 80 | 215 | 242-244 | 246 | 417 | 484 | 501 | 701 |
| Ancestral | L | D | D | LAL | R | K | E | N | A |
| B.1.351 | F | A | G | - - - | I | N | K | Y | V |
| SA-002 | F | A | G | - - - | R | N | K | Y | V |
| SA-006 | L | A | G | - - - | R | N | K | Y | V |
| SA-011 | L | A | G | - - - | R | N | K | Y | V |
| SA-012 | L | A | G | - - - | R | N | K | Y | V |
| SA-013 | L | A | G | - - - | R | N | K | Y | V |
| SA-017 | L | A | G | - - - | R | N | K | Y | V |
| SA-020** | L | A | G | - - - | R | N | K | Y | V |
| SA-024 | L | A | G | - - - | R | N | K | Y | V |
| SA-029** | L | A | D | - - - | I | N | K | Y | V |
| SA-037 | L | A | G | - - - | R | N | K | Y | V |
| SA-042 | F | A | G | - - - | R | N | K | Y | V |
| SA-043 | F | A | G | - - - | R | N | K | Y | V |
| SA-056 | F | A | G | - - - | R | N | K | Y | V |
| SA-070 | F | A | G | - - - | R | N | K | Y | V |
| SA-084 | F | A | G | - - - | R | N | K | Y | V |
| SA-094* | L | A | G | - - - | R | N | K | Y | V |
| SA-095 | F | A | G | - - - | R | N | K | Y | V |
| SA-100 | F | A | G | - - - | R | N | K | Y | V |
| SA-102 | F | A | G | - - - | R | N | K | Y | V |
| SA-003 | No sequence available | | | | | | | | |
| SA-016 | No sequence available | | | | | | | | |
| SA-052 | No sequence available | | | | | | | | |

**Fig. S1. Genomic sequencing confirmation of SARS-CoV-2 B.1.351 infection of COVID-19 second wave patients.** (**A**) A time-resolved maximum clade credibility phylogeny of 2621 global SARS-CoV-2 sequences. Sequences from South Africa (n=209) are denoted with tip points. The B.1.351 (501Y.V2) cluster is highlighted in red, and the genomes from this study (n=19) are shown in yellow, all falling in that cluster. (**B**) Spike sequence in patients recruited during the second wave, indicating amino acid changes. Blue shading corresponds to the ancestral strain (i.e. wild type, WT) amino acids and red shading to B.1.351 strain amino acids. (-) corresponds to amino acid deletions. $^{**}$: Patients exhibiting a detectable CD4 T cell response to the WT and B.1.351 peptide pools, $^{*}$: Patient exhibiting a detectable CD4 T cell response to the WT peptide pool.

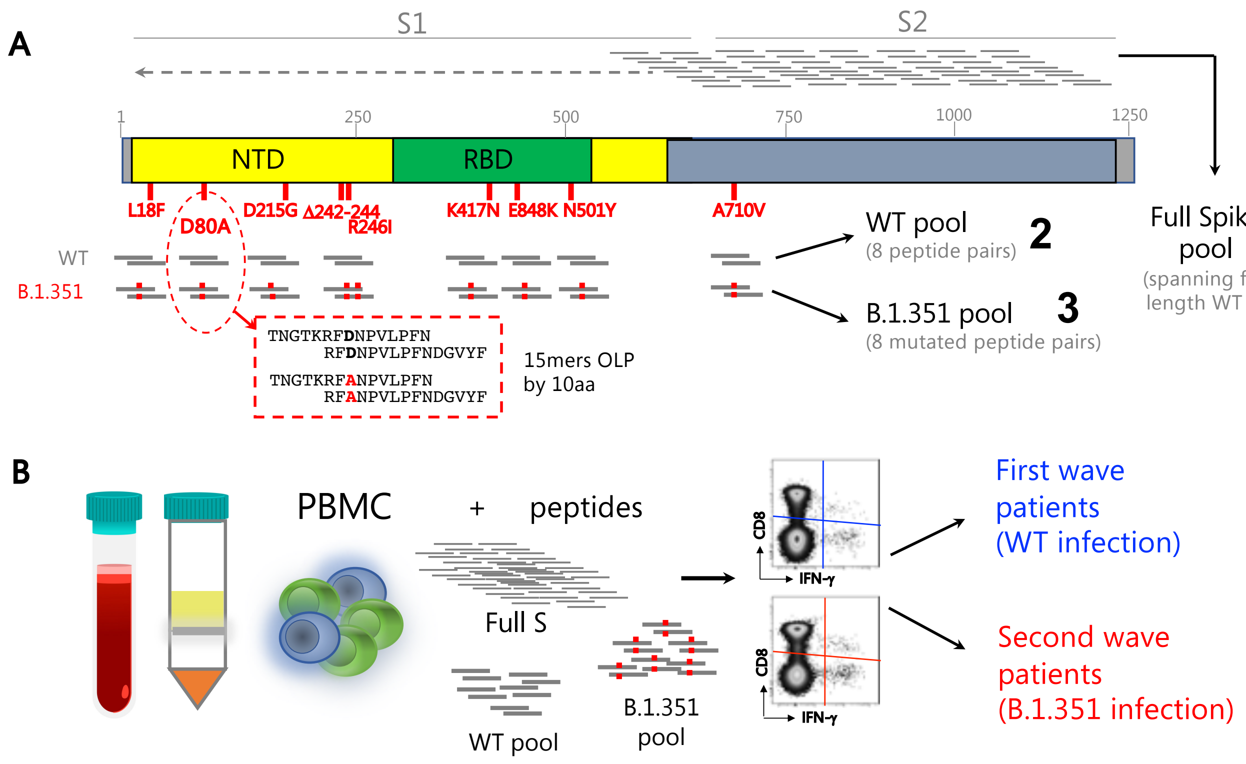


**Fig. S2. Graphical representation of study approach.** (**A**) SARS-CoV-2 spike protein is depicted, demonstrating the design of ancestral and B.1.351 variant peptides used in the immunological assays. (**B**) PBMC from patients recruited during the first or second wave of the pandemic in South Africa were stimulated with pools of peptides covering full spike, or smaller pools spanning only the regions mutated in B.1.351, with wild type (WT) and corresponding mutated versions.

**Table S1. Peptides included in the ancestral and B.1.351 peptide pools.**

| Mutations of interest | Strain | Position aa (start) | Position aa (stop) | Sequence | Previously described |
|---|---|---|---|---|---|
| L18F | ancestral | 6 | 20 | VLLPLVSSQCVNLTT | Mateus et al. |
| L18F | ancestral | 11 | 25 | VSSQCVNLTTRTQLP | Tarke et al. |
| D80A | ancestral | 73 | 87 | TNGTKRFDNPVLPFN | |
| D80A | ancestral | 78 | 92 | RFDNPVLPFNDGVYF | Tarke et al. |
| D215G | ancestral | 206 | 220 | KHTPINLVRDLPQGF | Tarke et al. |
| D215G | ancestral | 208 | 222 | TPINLVRDLPQGFSA | Peng et al. |
| D215G | ancestral | 211 | 225 | NLVRDLPQGFSALEP | Peng et al. |
| 242-244del/R246I | ancestral | 236 | 250 | TRFQTLLALHRSYLT | Mateus et al. |
| 242-244del/R246I | ancestral | 241 | 255 | LLALHRSYLTPGDSS | Mateus et al. |
| K417N | ancestral | 411 | 425 | APGQTGKIADYNYKL | |
| K417N | ancestral | 416 | 430 | GKIADYNYKLPDDFT | Mateus et al. |
| E484K | ancestral | 476 | 490 | GSTPCNGVEGFNCYF | |
| E484K | ancestral | 481 | 495 | NGVEGFNCYFPLQSY | |
| N501Y | ancestral | 492 | 506 | LQSYGFQPTNGVGYQ | |
| N501Y | ancestral | 496 | 510 | GFQPTNGVGYQPYRV | |
| A701V | ancestral | 701 | 715 | AENSVAYSNNSIAIP | Mateus et al. |
| L18F | B1.351 | 6 | 20 | VLLPLVSSQCVNFTT | |
| L18F | B1.351 | 11 | 25 | VSSQCVNFTTRTQLP | |
| D80A | B1.351 | 73 | 87 | TNGTKRFANPVLPFN | |
| D80A | B1.351 | 78 | 92 | RFANPVLPFNDGVYF | |
| D215G | B1.351 | 206 | 220 | KHTPINLVRGLPQGF | |
| D215G | B1.351 | 208 | 222 | TPINLVRGLPQGFSA | |
| D215G | B1.351 | 211 | 225 | NLVRGLPQGFSALEP | |
| 242-244del/R246I | B1.351 | 236 | 250 | TRFQTLHISYLTPGD | |
| 242-244del/R246I | B1.351 | 241 | 255 | LHISYLTPGDSSSGW | |
| K417N | B1.351 | 411 | 425 | APGQTGNIADYNYKL | |
| K417N | B1.351 | 416 | 430 | GNIADYNYKLPDDFT | |
| E484K | B1.351 | 476 | 490 | GSTPCNGVKGFNCYF | |
| E484K | B1.351 | 481 | 495 | NGVKGFNCYFPLQSY | |
| N501Y | B1.351 | 492 | 506 | LQSYGFQPTYGVGYQ | |
| N501Y | B1.351 | 496 | 510 | GFQPTYGVGYQPYRV | |
| A701V | B1.351 | 701 | 715 | VENSVAYSNNSIAIP | |

List and sequence of 15-mer peptides contained in the ancestral (WT) and B.1.351 peptide pools. References of previously described reactive peptides are provided in the last column. aa: amino acid. The site of mutation in B.1.351 is underlined in the peptide sequence.

**South African Cellular Immunology Network (SA-CIN) collaborators**

Fatima Abrahams[1], Frances Ayres[2], Elloise du Toit[3], Rene T. Goliath[1], Willem Hanekom[4], Diana Hardie[3], Nei-Yuan Hsiao[3], Henrik Kloverpris[4], Stephen Korsman[3], Francisco Lakay[1], Bronwen Lambson[2], Alasdair Leslie[4], Zanele Makhado[2], Moepeng Maseko[3], Donald Mhlanga[2], Michelle Naidoo[3], Zaza Ndhlovu[4], Amkele Ngomti[3], Brent Oosthuysen[2], Sheena Ruzive[1], Alex Sigal[4], Tamryn Smith[3], Ziyaad Valley-Omar[3], Dieter van der Westhuizen[5], Kennedy Zvinairo[1]

[1]Wellcome Centre for Infectious Diseases Research in Africa, University of Cape Town; Observatory 7925, South Africa
[2]National Institute for Communicable Diseases of the National Health Laboratory Services; Johannesburg, South Africa
[3]Division of Medical Virology, Department of Pathology, University of Cape Town; Observatory 7925, South Africa
[4]Africa Health Research Institute; Durban, South Africa
[5]Division of Chemical Pathology, Department of Pathology, University of Cape Town; Observatory 7925, South Africa
